# Study protocol: Assessing the association between corporate political influence and implementation of policies to tackle commercial determinants of non-communicable diseases: a cross-sectional analysis of 172 countries

**DOI:** 10.1101/2021.07.15.21260066

**Authors:** Luke N Allen, Simon Wigley, Hampus Holmer

## Abstract

**Objectives:** To assess the association between corporate political influence and implementation of WHO-recommended policies to constrain sales, marketing and consumption of tobacco, alcohol, and unhealthy foods.

**Design:** Cross-sectional analysis using national datasets from 2015, 2017, and 2020.

**Setting:** Global analysis of data from 172 of the 194 WHO Member States

**Main outcome measures:** We will use random effects multivariate regression to test the association between implementation status of 12 WHO-recommended tobacco, alcohol, and diet policies and *corporate political influence*, a metric that combines disclosure of campaign donations, public campaign finance, corporate campaign donations, legislature corrupt activities, disclosure by politicians, and executive oversight. We will control for GDP per capita, population aged >65 years, urbanization, level of democracy, continent, ethno-linguistic fractionalization, legal origin, Small Island Developing States, and Muslim population (to capture alcohol policy differences). We will include year dummies to address the possibility of a spurious relationship between the outcome variable and the independent variables of interests. For example, there may be an upward global trend in policy implementation that coincides with an upward global trend in in the regulation of lobbying and campaign finance.

**Ethics and dissemination:** As this study uses publicly available data, ethics approval is not required. The authors have no conflicts of interest to declare. Findings will be submitted to a peer-reviewed journal for publication in the academic literature. All data, code, and syntax will be made publicly available on GitHub.

## Background and rationale

Non-communicable diseases such as cancer, heart disease, diabetes and chronic obstructive pulmonary disease cause the majority of death and disability worldwide. This disease burden is largely preventable through a range of interventions, including a set of “Best Buy” policies endorsed by the 194 World Health Organization (WHO) Member States. Yet, many countries have not implemented these policies. It has been hypothesized that one reason may be the influence of corporations, particularly on those policies which seek to limit the consumption of unhealthy goods. Indeed, a growing body of work describes the myriad channels though which corporations seek to undermine effective public health measures in this way.^2–5^ Although case studies are plentiful, there has been a paucity of empirical research to quantify the association between corporate influence and policy implementation.

Among the WHO Best Buy policies are 12 policies that target tobacco, alcohol, foods high in fats and salt, child-focused junk food marketing, and marketing of breastmilk substitutes.^6^ These policies designed to tackle the commercial determinants of non-communicable diseases (NCDs) were first endorsed in 2013.^7^

WHO monitors the implementation of these commercial policies through regular NCD country capacity surveys, completed by national ministries of health. WHO has produced three global progress monitor reports – in 2015,^8^ 2017,^9^ and 2020^10^ – providing country-level assessments of whether each of the 12 commercial policies has been ‘fully implemented’, ‘partially implemented’, or ‘not implemented’.

This WHO data on commercial policy implementation provides a unique opportunity to examine whether indicators of corporate influence over policymaking processes are associated with implementation of key commercial policies, according to the three policy clusters delineated in Box 1.

### Box 1

**Globally-endorsed commercial policies from the WHO ‘best buys’**

Tobacco

- Tobacco tax
- Smoke free places
- Plain packaging and graphic warnings
- Tobacco advertising bans
- Tobacco mass media campaigns

Alcohol

- Alcohol sales restrictions
- Alcohol advertising bans
- Alcohol tax

Food

- Salt reduction
- Fat reduction
- Child marketing restrictions
- Restrictions on the marketing of breast milk substitutes

To elucidate the association between corporate influence and the implementation of commercial policies, in this exploratory analysis we will perform three sets of analyses:

1. To characterize implementation trends over time for tobacco, alcohol, and food-related policies using descriptive statistics, with sub-analysis by geographic region and income group.
2. To assess the association between implementation of commercial policies (aggregate score) and a newly developed measure of corporate political influence, controlling for a range of geopolitical variables using multivariatee regression. Sub-analyses will assess the association between:
  a. Country-level implementation of each of the three clusters (tobacco, alcohol, food) and corporate political influence.
  b. Country-level implementation of each individual policy and corporate political influence.
3. To identify countries with commercial policy implementation levels that are higher or lower than would be expected given their geopolitical characteristics; evaluated by creating a multivariate model and a modified Bland Altman chart.

We hypothesise that countries with the highest levels of corporate political influence will have the lowest levels of policy implementation.

In a secondary analysis we also aim to test whether the prevalence of smoking, alcohol use, hypertension, and adult and child obesity are respectively associated with implementation of tobacco, alcohol, salt, fat, and child marketing policies.

## Methods

This cross-sectional study will use observational data from a range of publicly available sources for the 194 WHO Member States for the years 2015, 2017, and 2020. All WHO Member States for which data were available will be included.

### Commercial policy implementation scores

Data on the implementation status of the 12 commercial policies (outlined in Box 1) for all 194 Member States will be extracted from the 2015, 2017, and 2020 WHO NCD Progress Monitor reports and transcribed into a csv spreadsheet. Data will be double-checked by two authors. Full descriptions of each policy are available in Appendix 1. Following the approach of WHO and Allen et al.^11^ we will construct policy scores for each country, according 1 point for each fully implemented policy, 0.5 points for each partially implemented policy, and 0 points for non-implemented policies and those for which no data are available. We will construct an overall aggregate score for each country, ranging from 0-12, as well as policy cluster scores for tobacco (range 0-5), alcohol (range 0-3) and food (range 0-4).

### Commercial political influence

We aim to assess whether direct commercial political influence – i.e. payments to politicians and their parties - (independent variable) is associated with implementation of commercial NCD policies (dependent variable). Whilst there are myriad examples of corporate actors using their financial clout to undermine NCD regulations,^12^ it is important to note that policymaking is a complex process and corporations do not universally seek to undermine effective NCD policies.

This analysis will focus on a narrow conceptual space concerning whether corporate actors wield outsized financial influence over policymakers, meaning that the arguments and lobbying efforts of other non-commercial actors (such as public health advocates) – are marginalized.^13^ The political science and global health literature consistently identify four regulatory areas in this space:^2,5,14–22^

1. *Campaign financing*: Are there limits on campaign donations from companies and/or a requirement to publicly disclose the source and amount of donations whether there are limits on campaign donations from companies and/or a requirement to publicly disclose the source and amount of those donations?
2. *Business and financial interests of politicians*: Are there mandatory public disclosures of politicians’ financial and business interests?
3. *Lobbying transparency*: Are there mandatory public disclosures of lobbyists activities?
4. *Enforcement*: Is there an independent administrative or judicial body has the capacity to enforce the above-listed financing limits and disclosure requirements?

As far as we are aware, there is not a single globally comparable indicator that combines these four domains to quantify the level of *corporate political influence* in each country. As such, we performed a literature review to identify the most robust, globally comparable, and conceptually aligned metric to use as the independent variable, reported in Appendix 2. The closest - Lima and Galea’s corporate permeation index (CPI) - includes a wide variety of input variables, meaning that the scope of that metric extends well beyond the ability of corporations to directly influence the policymaking process. Rather, CPI captures “the extent to which corporations are embedded in the political, legal, social, economic and cultural fabric of a given society”. Furthermore, their CPI metric only covers 146 countries.^23^ Whilst there was not a single composite indicator that captured commercial political influence, our review did identify six individual proxies that were well aligned with three of the four regulatory areas (we were unable to identity an indicator of lobbying transparency with sufficient country coverage). These items all conceptually map to the political-commercial nexus, have strong internal and external validity, and cover 172 countries (22 microstates are excluded, see Appendix 3 for list).

#### Box 2

**Indicators that capture different aspects of corporate political influence**

1. ***Disclosure of campaign donations***: *Are there disclosure requirements for donations to national election campaigns? (Source: V-Dem Dataset v11*.*1)*
2. ***Public campaign finance***: *Is significant public financing available for parties’ and/or candidates’ campaigns for national office? (Source: V-Dem Dataset v11*.*1)*
3. ***Corporate campaign donations***: *Is there a ban on donations from domestic or foreign interests to political parties or candidates? (Source: IDEA. Political Finance Database, 2020 update)*
4. ***Disclosure by politicians***: *Do the law or regulations of the country require politicians to provide either financial and/or business interests disclosures and are the disclosures publicly available? (Source: Djankov et al, 2010* ^24^)
5. ***Legislature corrupt activities***: *Do members of the legislature abuse their position for financial gain? (Source: V-Dem Dataset v11*.*1)*
6. ***Executive oversight***: *If executive branch officials were engaged in unconstitutional, illegal, or unethical activity, how likely is it that a body other than the legislature, such as a comptroller general, general prosecutor, or ombudsman, would question or investigate them and issue an unfavorable decision or report? (Source: V-Dem Dataset v11*.*1)*

Building on the work of Lima and Galea, we will use structural equation modelling (with full information maximum likelihood) in Stata to identify the latent factor underlying the six input indicators listed in Box 2. This will enable us to create a new *Corporate Political Influence Index* (CPII) that focuses on the interaction between politicians and commercial actors. Whilst there are no direct indicators for lobbying currently available, it is reasonable to expect that the latent factor (the single underlying factor picked out by the factor analysis) behind the six included variables will capture lobbying activities. Having identified a single underlying factor, we will generate an index score for each country, ranging from zero (lowest level of corporate political influence) to 100 (highest).

### Control variables

In assessing the association between CPII and commercial policy implementation, we will control for the following economic, cultural, historical, geographic, and population factors: GDP per capita, population aged >65 years, urbanization, level of democracy, continent, ethno-linguistic fractionalization, legal origin, Small Island Developing States, and Muslim population (to capture alcohol policy differences) – control variables derived from earlier work on international NCD policy implementation.^11^ We will include year dummies to address global trends in terms of the outcome variable and the independent variables of interest. For example, there may be an upward global trend in policy implementation that coincides with an upward global trend in in the regulation of lobbying and campaign finance.

### Statistical analyses

We will use descriptive statistics to characterize implementation trends over time for the commercial policies. We will present mean implementation scores for each WHO geographic region and World Bank income group.

We will perform the following three random effects regression analyses:

#### Ia: Aggregate policy score

Total commercial policies (aggregate score for all 12 policies) regressed on CPII.

#### Ib: Policy clusters

Each commercial policy cluster (tobacco, alcohol, and food) separately regressed on CPII.

#### Ic: Individual policies

All 12 individual commercial policies separately regressed on CPII.

We will use random effects GLS regressions to capture between-country effects and within-country effects, using Stata’s xtreg, re command. We will perform each regression with and without controls.

### Identification of outliers

We will use the results from Ia and Ib to construct prediction-based modified Bland-Altman plots for 2020, plotting each country’s WHO-ascertained policy implementation score on the x axis, and predicted score on the y axis, based on the regression equation. We will set 95% limits of agreement to identify over- and under-performing countries.

### Additional model

#### Risk factor prevalence and policy implementation

We will use the random effects GLS model to test whether commercial policy implementation is associated with the prevalence of the following risk factors at the national level:

i. Tobacco cluster aggregate score vs Total smoking prevalence (ages 15+).^25^
ii. Alcohol cluster aggregate score vs Alcohol consumption per capita (ages 15+).^26^
iii. Salt reduction policy score vs Hypertension prevalence (ages 18+).^27,28^
iv. Fat reduction policy score vs Prevalence of BMI >30 (ages 18+).^29,30^
v. Child food marketing policy score vs Prevalence of BMI >30 (ages <18).^29,30^

### Sensitivity analyses and robustness checks

We will repeat the three regression models using Lima and Galea’s Corporate Permeation Index^23^, a version of CPII that includes the registration of lobbying activities (only available for 127 countries), and a further version of CPII that drops Djankov’s ‘disclosures by politicians’ data (only available for 2010). We will also repeat the three regression models and the additional risk factor prevalence regression using multiple imputation to address missing data, using Stata’s mi impute mvn and mi est commands.

We will repeat the regression models including level of corruption as a control variable as it is a potential confounders for CPII. We will use the *Political Corruption Index* from the V-Dem dataset, version 11.1. We will perform multiplicity tests for all regression models using Stats’s wyoung module.

We will produce variable and coefficient matrices for regression model Ia in order to check for collinearity, using Stata. Finally, we will perform the Robust Hausman test for random vs. fixed effects using Stata’s rhausman module.

### Data management and statistical principles

All raw data used in the study will be uploaded to GitHub and made publicly available. All analyses will be performed on Stata version 14.1 and R version 4.1.0. We will use a 0.05 level of statistical significance, cluster-robust standard errors, and report 95% confidence intervals. We will follow the statistical analysis plan in Appendix 4, which was developed in line with the DEBATE reporting guidelines for observational studies.^31^ We will upload our syntax to: https://github.com/drlukeallen/CDOH-policy-implementation

## Data Availability

All data, code, and syntax will be made publicly available on GitHub.

https://github.com/drlukeallen/CDOH-policy-implementation

## Ethics and dissemination

As this study uses publicly available data, ethics approval is not required. Full access to all data used in the study will be provided via GitHub. The authors have no conflicts of interest to declare.

Findings will be submitted to a peer-reviewed journal for publication in the academic literature.

## Appendix 1 Full definitions of the commercial policies included in the WHO NCD progress monitors

### Tobacco policy cluster

- 5a: Member State has implemented measures to reduce affordability by increasing excise taxes and prices on tobacco products
- 5b: Member State has implemented measures to eliminate exposure to second-hand tobacco smoke in all indoor workplaces, public places and public transport
- 5c: Member State has implemented plain/ standardized packaging and/or large graphic health warnings on all tobacco packages
- 5d: Member State has enacted and enforced comprehensive bans on tobacco advertising, promotion and sponsorship
- 5e: (2017 and 2020 only) Member State has implemented effective mass media campaigns that educate the public about the harms of smoking/tobacco use and second-hand smoke

### Alcohol policy cluster

- 6a: Member State has enacted and enforced restrictions on the physical availability of retailed alcohol (via reduced hours of sale)
- 6b: Member State has enacted and enforced bans or comprehensive restrictions on exposure to alcohol advertising (across multiple types of media)
- 6c: Member State has increased excise taxes on alcoholic beverages

### Food policy cluster

- 7a: Member State has adopted national policies to reduce population salt/sodium consumption
- 7b: Member State adopted national policies that limit saturated fatty acids and virtually eliminate industrially produced trans-fatty acids in the food supply
- 7c: Member State has implemented the WHO set of recommendations on marketing of foods and non-alcoholic beverages to children
- 7d: Member State has legislation/regulations fully implementing the International Code of Marketing of Breast-milk Substitutes

## Appendix 2 Literature review: Proxies for the commercial-political nexus

### Background

We conducted a literature review to identify indicators that can be used to assess the extent to which commercial actors are able to directly influence the policy-making process. The central issue is not whether commercial interests can influence policy, but whether they have an outsized influence over policy-making due to their financial clout.^13^ The latter would grant them an advantage over non-commercial interest groups, including those advocating for public health measures to constrain the sale and marketing of unhealthy products. Much depends, therefore, on whether there are formal restrictions on the use of corporate resources to directly influence policy-makers.

The legal and political science literature on political financing and the role of interest groups is extensive.^14–18,32^ Similarly a large public health literature has emerged on the use of corporate resources to influence health policy.^2,5,19–22^ Based on these two literatures we identified four regulatory areas that affect the ability of commercial interests to use their greater financial resources to directly influence politicians and political parties and, thereby, the policy-making process:

1. *Campaign financing*: whether there are limits on campaign donations from companies and/or a requirement to publicly disclose the source and amount of those donations.
2. *Business and financial interests of politicians*: whether the financial and business interests of politicians must be publicly disclosed.
3. *Lobbying transparency*: whether the activities of lobbyists must be publicly disclosed.
4. *Enforcement*: whether an independent administrative or judicial body (e.g. electoral monitoring board, general prosecutor, etc.) has the capacity to enforce financing limits and disclosure requirements (i.e. 1-3).

It is important to note that most of the channels through which corporations can use their resources to influence policy-making do not involve corrupt incentives in the traditional sense (i.e. the offering of material incentives that personally benefit politicians). Lobbying activities and campaign donations typically do not provide material benefits to individual politicians. Campaign donations enhance the electoral chances of politicians and political parties, but individual politicians may only materially benefit if they embezzle those donations.^33^ Nevertheless, corrupt incentives remain one way in which commercial interests can gain unequal influence over policy. This is covered by the second regulatory area above.

We set aside the ability of corporations to *indirectly* influence policy-makers by shaping consumer preferences (via, for example, product advertising and media ownership) and, thereby, the policies that citizens support.^19^ Instead we focus on the ability of corporations to *directly* influence the decisions of policy-makers.

We aimed to construct a robust overall indicator that captures the level of regulation in each of these four areas. To achieve this, we sought to identify individual indicators for each regulatory area that could then be combined via factor analysis to form a *de novo* composite indicator of corporate political influence.

### Method

#### Search strategy

To identify indicators of corporate influence over political decision-making aligned with the four domains listed above, we searched the following global political data collections on July 7^th^ 2021: Varieties of Democracy (V-Dem) data set,^34^ Quality of Government (QoG) standard data set,^35^ and Institute for Democracy and Electoral Assistance’s (IDEA) Political Finance Database.^36^ In addition, we searched the first 1,000 records of Google Scholar using the following broad search terms; (“disclosure” OR “transparency”) AND (“politician” OR “lobbying” OR “donation” OR “campaign financing”) AND “data”. We limited the search to the years 2005-2021.

##### Inclusion criteria

We included metrics that met the following criteria:

- Conceptual alignment with the four regulatory areas noted above
- Global coverage: data available for an arbitrary threshold of >85% (>165) WHO Member States
- Data gathering methodology is publicly available and complete
- Data is comparable between countries

Two authors (SW and LA) independently reviewed each potential indicator for alignment with the inclusion criteria. Disagreements were resolved by discussion and - where necessary – arbitration by the third author (HH). The rationale for inclusion or exclusion was documented for each indicator.

### Findings

Table 1 presents the set of relevant indicators that have global coverage, as well as the rationale for inclusion and the quality of the data gathering process in each case.

**Table 1.**

| Indicator | Description | Rationale | Source | Country coverage | Data gathering methodology publicly available and complete? | Data gathering method | Comparable between countries? | Scale | Years | Include/exclude |
| --- | --- | --- | --- | --- | --- | --- | --- | --- | --- | --- |
| **Campaign finance** |  |  |  |  |  |  |  |  |  |  |
| **Disclosure of campaign donations** | To what extent are there disclosure requirements for donations to national election campaigns? | Absence of transparency increases the opportunity for commercial interests to influence politicians and political parties and, thereby, policy. | V-Dem Dataset v11.1<sup>34</sup> [variable name: v2eldonate] | 172 | yes | Coded by multiple country experts and converted to interval scale using Bayesian item response theory measurement model. | yes | Interval scale where the lowest score indicates the least disclosure and the highest score indicates the most disclosure. | 2015, 2017, 2019 | Meets inclusion criteria - include |
| **Public campaign finance** | To what extent is significant public financing available for parties' and/or candidates' campaigns for national office? | Greater public financing ameliorates the influence of large private donors on politicians and political parties and, thereby, policy-making. | V-Dem Dataset v11.1<sup>34</sup> (variable name: v2elpubfin) | 172 | yes | Coded by multiple country experts and converted to interval scale using Bayesian item response theory measurement model. | yes | Interval scale where the lowest score indicates the least public financing and highest score indicates the most public financing. | 2015, 2017, 2019 | Less direct, but still meets inclusion criteria - include |
| **Corporate campaign donations** | Is there a ban on donations from domestic or foreign interests to political parties or candidates? | Ban reduces the opportunity for commercial interests to influence politicians and political parties and, thereby, policy. | IDEA (2021)<sup>36</sup> | 180 | yes | Expert coded based on laws, regulations, and reports.<br>Aggregate score is the average of questions 1-4 listed under 'Bans and limits on private income' in the IDEA database:<br>1. Is there a ban on donations from foreign interests to political parties?<br>2. Is there a ban on donations from foreign interests to candidates?<br>3. Is there a ban on corporate donations to political parties?<br>4. Is there a ban on corporate donations to candidates? | yes | Interval scale ranging from 0 to 1, where 0 represents no restrictions on donations from corporations and 1 represents the most restrictions on donations from corporations. | 2019 | Discussion around whether the foreign element for this composite marker excludes it. We decided to include on the basis that most foreign donations are likely to come from corporate actors. |
| **Disclosure of interests** |  |  |  |  |  |  |  |  |  |  |
| **Disclosure by politicians** | Do the law or regulations of the country require politicians to provide either financial and/or business interests disclosures and are the disclosures publicly available? | Absence of transparency increases the opportunity for commercial interests to influence politicians and political parties and, thereby, policy. | Djankov et al (2010)<sup>24</sup> [variable names: disc_req and ft_pubprac_all] | 175 | yes | Expert coded based on laws, regulations, and information requests.<br>Score is the sum of disc_req and ft_pubprac_all | yes | Ordinal scale:<br>0 Disclosure not required and not publicly available<br>1 Disclosure required or publicly available<br>1.5 Disclosure required but financial or business disclosures not publicly available.<br>2 Disclosure required and publicly available | 2010 | Data are quite old; however, this is a slow-moving domain. As such we decided to include, but run a sensitivity analysis that drops this variable from the latent factor analysis. |
| **Legislature corrupt activities** | To what extent do members of the legislature abuse their position for financial gain? | Willingness of politicians to accept material incentives for personal gain increases the opportunity for commercial interests to influence policy. | V-Dem Dataset v11.1<sup>34</sup> [variable name: v2lgcrprt] | 172 | yes | Coded by multiple country experts and converted to interval scale using Bayesian item response theory measurement model. | yes | Interval scale where the lowest score indicates the most corruption and the highest score indicates the least corruption. | 2015, 2017, 2019 | We noted that we are interested in licit and illicit financial influence, not just corruption. However this indicator is likely to helpfully capture non-disclosures, and is a proxy for opportunities for influencing politicians. We agreed to include. |
| **Lobbying transparency** |  |  |  |  |  |  |  |  |  |  |
| **Registration of lobbying activities** | Are lobbyists for commercial interests required to register their identity and the company they represent? | Absence of transparency increases the opportunity for commercial interests to influence politicians and political parties and, thereby, policy. | IDEA (2021)<sup>36</sup>; OECD (2021)<sup>13</sup>; Chari et al (2019)<sup>37</sup> | 127 | yes | Expert coded based on laws and regulations.<br>This variables was constructed based on the three sources noted. Each country was sorted according to the following four categories: (1) no disclosure requirements, (2) voluntary registration of lobbying activities, (3) some lobbying activities must be registered, (4) most lobbying activities must be registered. The binary variable takes the value of 1 if some or most activities must be registered, otherwise 0. | yes | Dichotomous variable which takes the value of zero if there is no requirement to register lobbying activities, otherwise 1. | 2019 | Because data is not available for a sufficient number of countries we did not use this indicator during the construction of CPII. We assume that the latent variable identified using factor analysis will also capture this regulatory area - countries with demanding disclosure requirements for campaign donations and the financial/business interests of politicians are more likely to have disclosure requirement for lobbying activities, and vice versa. By way of robustness we will test whether our regression results are sensitive to the inclusion of this variable in the factor analysis. |
| **Enforcement** |  |  |  |  |  |  |  |  |  |  |
| **Executive oversight** | If executive branch officials were engaged in unconstitutional, illegal, or unethical activity, how likely is it that a body other than the legislature, such as a comptroller general, general prosecutor, or ombudsman, would question or investigate them and issue an unfavorable decision or report? | Indicates the likelihood that bans and disclosure requirements are enforced in practice. | V-Dem Dataset v11.1<sup>34</sup> [variable name: v2lgotovst] | 172 | yes | Coded by multiple country experts and converted to interval scale using Bayesian item response theory measurement model. | yes | Interval scale where the lowest score indicates the least oversight and the highest score indicates the most oversight. | 2015, 2017, 2019 | Meets the inclusion criteria - include. |

We identified a total of six indicators that were methodologically robust, available for >80% of countries, internationally comparable, and conceptually aligned with the ability of commercial actors to influence policymakers. As we were only able to find lobbying transparency data for 127 countries that input indicator was set aside. However, it is reasonable to expect that the other three regulatory areas are correlated with the level of transparency in terms of lobbying activities. That is, polities that require the disclosure of campaign donations and business/financial interests are more likely to require disclosure by lobbyists. For the sake of robustness we will check whether our regression results are sensitive to the inclusion of the lobbying transparency indicator during the construction of the CPII.

## Appendix 3 List of 22 microstates not covered by the V-Dem data

- Andorra
- Antigua and Barbuda
- Bahamas
- Belize
- Brunei Darussalam
- Cook Islands
- Dominica
- Micronesia
- Grenada
- Kiribati
- Saint Kitts and Nevis
- Saint Lucia
- Monaco
- Marshall Islands
- Niue
- Nauru
- Palau
- San Marino
- Tonga
- Tuvalu
- Saint Vincent and the Grenadines
- Samoa

## Appendix 4 Statistical analysis plan

### Administrative information

1. Title: Implementation of policies to tackle the commercial determinants of non-communicable diseases from 2015 to 2020: a cross-sectional analysis of 172 countries exploratory analysis
2. SAP version: Version 1.0 (July 1 2021)
3. SAP Revisions:
4. Roles and responsibility:
  i. Person writing SAP: Simon Wigley and Luke Allen
  ii. Senior statistician responsible: Simon Wigley
  iii. Chief investigator: Luke Allen

### Introduction

5. Background and rationale: A growing body of work describes the myriad channels though which corporations seek to undermine effective public health measures to constrain the sale and marketing of unhealthy commodities. [1–6] Although case studies are plentiful, there has been a paucity of empirical research to quantify the association between corporate influence and policy implementation. All 194 World Health Organization (WHO) Member States have endorsed a set of 12 policies that target tobacco, alcohol, foods high in fats and salt, child-focused junk food marketing, and marketing of breastmilk substitutes. These policies designed to tackle the ‘commercial determinants’ of non-communicable diseases (NCDs) were first endorsed in 2013. WHO monitors the implementation of these commercial policies through regular NCD country capacity surveys, completed by national ministries of health. WHO has produced three global progress monitor reports – in 2015, 2017, and 2020 [7–9] – providing country-level assessments of whether each of the 12 commercial policies has been ‘fully implemented’, ‘partially implemented’, or ‘not implemented’. This WHO data on commercial policy implementation provides a unique opportunity to examine whether indicators of corporate influence over policymaking processes are associated with implementation of key commercial policies, according to the three policy clusters delineated in Box 1.

#### Box 1

Commercial policies in three ‘policy clusters’

***Tobacco***

- Tobacco tax
- Smoke free places
- Tobacco packaging
- Tobacco advertising bans
- Tobacco mass media

***Alcohol***

- Alcohol sales restrictions
- Alcohol advertising bans
- Alcohol tax

**Food**

- Salt reduction
- Fat reduction
- Child marketing restrictions
- Marketing of breast milk substitutes

6. Objectives In this exploratory analysis we aimed to provide a summary of international implementation trends over time, develop a composite corporate political influence index (CPII), and perform three sets of regression analyses assess the association between:
  1. Overall country-level implementation of all 12 commercial policies (aggregate score) and CPII.
  2. Country-level implementation of each of the three clusters and CPII.
  3. Country-level implementation of each individual policy and CPII.
  4. Country level prevalence of risk factors (tobacco use, alcohol use, hypertension, obesity, and child obesity) and each of the respective policies.

We hypothesise that countries with higher levels of corporate political influence will have the lowest levels of policy implementation.

### Study methods

7. Study design: cross-sectional study based on observational data

### Statistical principles

8. Confidence intervals and p-values.
  i. Level of statistical significance: 0.05
  ii. Confidence interval to be reported: 95%
  iii. Adjustment for multiplicity: see 32 below

### Study sample

9. 172-194 sovereign states for the years 2015, 2017, and 2019.

### Data gathering and construction

10. Extract policy scores from NCD Progress Monitors for 2015, 2017 and 2020. [7– 9]
11. Construct composite scores for all 12 commercial policies and each commercial policy cluster (tobacco, alcohol and food policies).
12. Assemble risk factors from various sources: adult obesity, child obesity, hypertension, alcohol consumption, and smoking prevalence.
13. Assemble control variables from various sources: GDP per capita, % urban population, % population aged 65+, level of democracy, % Muslim, legal origin, ethno-linguistic fractionalization, Small Island Developing States, and continent.
14. Construct ‘Corporate Political Influence’ index using latent factor analysis, using structural equation modelling in Stata with FIML.
15. Exclude variables that encompass less than 165 of 194 countries.
16. Log transform variables that are right-skewed.
17. Use data from 10-16 to construct a panel for the years 2015, 2017, and 2019 for 172-194 countries.

### Model specification

18. Random effects GLS regressions: captures between country effects and within country effects
19. Confounding covariates: include control variables to address economic, cultural/ historical, geographic, and population factors that may be driving the results.
20. Time trends: include year dummies to address the possibility of a spurious relationship between the outcome variable and the independent variables of interests.
21. Standard errors: cluster-robust

### Regression analyses

22. Random effects regression I: total commercial policies (aggregate score for all 12 policies) regressed on CPII (with and without control variables) [using Stata’s xtreg, re command].
23. Random effects regression II: each commercial policy cluster (tobacco, alcohol, and food) separately regressed on CPII (with and without control variables) [using Stata’s xtreg, re command]
24. Random effects regressions III: All 12 individual commercial policies separately regressed on CPII (with and without control variables) [using Stata’s xtreg, re command]
25. Construct prediction-based Bland-Altman plots for 2019 using the results from 22 and 23.

### Additional analysis

26. Random effects regressions IV: tobacco, alcohol, fat, child food marketing, and salt policies separately regressed on corresponding risk factors (with and without control variables) [using Stata’s xtreg, re command]
27. Interaction model: examine whether the association between democracy and commercial policies is conditioned by level of CPII. [using Interflex command in R][10]

### Robustness/ sensitivity checks

28. Repeat 22, 23, 24, and 27 using Lima and Galea’s [11] Corporate Permeation Index.
29. Repeat 22, 23, 24, and 27 using CPII constructed:
  i. Without disclosure by politicians (because data for that variable is only available for year 2010)
  ii. With registration of lobbying activities (which is only available for 127 countries)
30. Repeat 22, 23, 24, and 26 using multiple imputation to address missing data [using Stata’s mi impute mvn and mi est commands]
31. Repeat 22, 23, 24, and 27 including level of corruption as a control variable (potential confounder for CPII).
32. Collinearity checks: Variable and coefficient correlation matrices for 22.
33. Multiplicity tests for regressions [using Stata’s wyoung module]
34. Robust Hausman test for random vs. fixed effects [using Stata’s rhausman module]

### Statistical software

35. Stata version 14.1 and R version 4.1.0

## References

1 Chan A-W, Tetzlaff JM, Altman DG, et al. SPIRIT 2013 statement: defining standard protocol items for clinical trials. Ann Intern Med 2013; 158: 200–7.

2 McKee M, Stuckler D. Revisiting the Corporate and Commercial Determinants of Health. Am J Public Health 2018; 108: 1167–70.

3 Hawkins B, Holden C. A Corporate Veto on Health Policy? Global Constitutionalism and Investor-State Dispute Settlement. J Health Polit Policy Law 2016; 41: 969–95.

4 Miller D, Harkins C. Corporate strategy, corporate capture: Food and alcohol industry lobbying and public health. Crit Soc Policy 2010; 30: 564–89.

5 Allen LN. Commercial Determinants of Global Health. In: Haring R, Kickbusch I, Ganten D, Moeti M, eds. Handbook of Global Health. Cham: Springer International Publishing, 2020: 1–37.

6 World Health Organization. Tackling NCDs: ‘best buys’ and other recommended interventions for the prevention and control of noncommunicable diseases. Updated (2017) appendix 3 of the global action plan for the prevention and control of noncommunicable diseases 2013-2020. Endorsed at the 70th World Health Assembly, 2017. Geneva: World Health Organization, 2017 https://apps.who.int/iris/handle/10665/259232 (accessed July 9, 2021).

7 World Health Assembly. Resolution WHA66/9. Draft action plan for the prevention and control of non-communicable diseases 2013–2020. 2013. https://apps.who.int/gb/e/e_wha66.html (accessed July 12, 2021).

8 Noncommunicable Diseases Progress Monitor 2015. Geneva: World Health Organization, 2015 https://www.who.int/publications-detail-redirect/noncommunicable-diseases-progress-monitor-2015 (accessed July 1, 2021).

9 Noncommunicable Diseases Progress Monitor 2017. Geneva: World Health Organization, 2017 https://www.who.int/publications-detail-redirect/9789241513029 (accessed July 1, 2021).

10 Noncommunicable Diseases Progress Monitor 2020. Geneva: World Health Organization, 2020 https://www.who.int/publications-detail-redirect/ncd-progress-monitor-2020 (accessed July 1, 2021).

11 Allen LN, Nicholson BD, Yeung BYT, Goiana-da-Silva F. Implementation of non-communicable disease policies: a geopolitical analysis of 151 countries. Lancet Glob Health 2020; 8: e50–8.

12 Allen LN. Commercial Determinants of Global Health. In: Haring R, Kickbusch I, Ganten D, Moeti M, eds. Handbook of Global Health. Cham: Springer International Publishing, 2020: 1–37.

13 OECD. Lobbying in the 21st Century: Transparency, Integrity and Access. OECD, 2021 DOI:10.1787/c6d8eff8-en.

14 de Figueiredo JM, Richter BK. Advancing the Empirical Research on Lobbying. Annu Rev Polit Sci 2014; 17: 163–85.

15 Falguera E, Jones S, Ohman M, International Institute for Democracy and Electoral Assistance, editors. Funding of political parties and election campaigns: a handbook on political finance. Stockholm: IDEA, 2014.

16 Briffault R. Lobbying and Campaign Finance: Separate and Together Symposium: The Law of Lobbying. Stanf Law Policy Rev 2008; 19: 105–29.

17 Gilens M, Patterson S, Haines P. Campaign Finance Regulations and Public Policy. Am Polit Sci Rev forthcoming. DOI:10.1017/S0003055421000149.

18 Crepaz M. Why do we have lobbying rules? Investigating the introduction of lobbying laws in EU and OECD member states. Interest Groups Advocacy 2017; 6: 231–52.

19 Madureira Lima J, Galea S. Corporate practices and health: a framework and mechanisms. Glob Health 2018; 14: 21.

20 Hanefeld J, Reeves A, Brown C, Östlin P. Achieving health equity: democracy matters. The Lancet 2019; 394: 1600–1.

21 Wiist WH. Citizens United, Public Health, and Democracy: The Supreme Court Ruling, Its Implications, and Proposed Action. Am J Public Health 2011; 101: 1172–9.

22 Nestle M. Food Politics: How the Food Industry Influences Nutrition, and Health, Revised and Expanded Edition, Revised edition. Berkeley: University of California Press, 2013.

23 Madureira Lima J, Galea S. The Corporate Permeation Index – A tool to study the macrosocial determinants of Non-Communicable Disease. SSM - Popul Health 2019; 7: 100361.

24 Djankov S, La Porta R, Lopez-de-Silanes F, Shleifer A. Disclosure by Politicians. Am Econ J Appl Econ 2010; 2: 179–209.

25 Reitsma MB, Kendrick PJ, Ababneh E, et al. Spatial, temporal, and demographic patterns in prevalence of smoking tobacco use and attributable disease burden in 204 countries and territories, 1990–2019: a systematic analysis from the Global Burden of Disease Study 2019. The Lancet 2021; 397: 2337–60.

26 World Bank. World Development Indicators 2021. Washington D.C.: World Bank, 2021 https://databank.worldbank.org/source/world-development-indicators (accessed July 7, 2021).

27 NCD RisC. Blood Pressure. Evol. Blood Press. Time. https://ncdrisc.org/data-downloads-blood-pressure.html (accessed July 12, 2021).

28 Zhou B, Bentham J, Cesare MD, et al. Worldwide trends in blood pressure from 1975 to 2015: a pooled analysis of 1479 population-based measurement studies with 19·1 million participants. The Lancet 2017; 389: 37–55.

29 NCD-RisC. National Body Mass Index. Evol. BMI Time. https://ncdrisc.org/data-downloads-adiposity.html (accessed July 12, 2021).

30 Abarca-Gómez L, Abdeen ZA, Hamid ZA, et al. Worldwide trends in body-mass index, underweight, overweight, and obesity from 1975 to 2016: a pooled analysis of 2416 population-based measurement studies in 128·9 million children, adolescents, and adults. The Lancet 2017; 390: 2627–42.

31 Hiemstra B, Keus F, Wetterslev J, Gluud C, van der Horst ICC. DEBATE-statistical analysis plans for observational studies. BMC Med Res Methodol 2019; 19: 233.

32 Erne R. Interest Groups. In: Caramani D, ed. Comparative politics, Fifth edition. Oxford: Oxford University Press, 2020: 252–66.

33 Thompson DF. Mediated Corruption: The Case of the Keating Five. Am Polit Sci Rev 1993; 87: 369–81.

34 Coppedge M, et al. V-Dem [Country–Year/Country–Date] Dataset v11.1. 2021. DOI:10.23696/VDEMDS21.

35 Teorell J, Dahlberg S, Sundström A, et al. QoG Standard Dataset 2021. 2021. DOI:10.18157/QOGSTDJAN21.

36 IDEA. Political Finance Database. 2020 update. https://www.idea.int/data-tools/data/political-finance-database (accessed May 5, 2021).

37 Chari RS, Hogan J, Murphy G, Crepaz M. Regulating lobbying: a global comparison, 2nd edition. Manchester: Manchester University Press, 2019.

## References for SAP

1 McKee M, Stuckler D. Revisiting the Corporate and Commercial Determinants of Health. Am J Public Health 2018;108:1167–70.

2 Gilmore AB, Fooks G, Drope J, et al. Exposing and addressing tobacco industry conduct in low-income and middle-income countries. The Lancet 2015;385:1029–43.

3 Miller D, Harkins C. Corporate strategy, corporate capture: Food and alcohol industry lobbying and public health. Crit Soc Policy 2010;30:564–89.

4 Hawkins B, Holden C. A Corporate Veto on Health Policy? Global Constitutionalism and Investor-State Dispute Settlement. J Health Polit Policy Law 2016;41:969–95.

5 Allen LN. Commercial Determinants of Global Health. In: Haring R, Kickbusch I, Ganten D, et al., eds. Handbook of Global Health. Cham: : Springer International Publishing 2020. 1– 37.

6 Corporate Accountability. Partnership for an unhealthy planet: How big business interferes with global health policy and science. Corporate Accountability 2020. https://www.corporateaccountability.org/resources/partnership-for-an-unhealthy-planet/ (accessed 25 Sep 2020).

7 Noncommunicable Diseases Progress Monitor 2015. Geneva: : World Health Organization 2015. https://www.who.int/publications-detail-redirect/noncommunicable-diseases-progress-monitor-2015 (accessed 1 Jul 2021).

8 Noncommunicable Diseases Progress Monitor 2017. Geneva: : World Health Organization 2017. https://www.who.int/publications-detail-redirect/9789241513029 (accessed 1 Jul 2021).

9 Noncommunicable Diseases Progress Monitor 2020. Geneva: : World Health Organization 2020. https://www.who.int/publications-detail-redirect/ncd-progress-monitor-2020 (accessed 1 Jul 2021).

10 Hainmueller J, Mummolo J, Xu Y. How Much Should We Trust Estimates from Multiplicative Interaction Models? Simple Tools to Improve Empirical Practice. Polit Anal 2019;27:163–92.

11 Madureira Lima J, Galea S. The Corporate Permeation Index – A tool to study the macrosocial determinants of Non-Communicable Disease. SSM - Popul Health 2019;7:100361.

